# ProtFI, an efficient frailty-trained proteomics-based biomarker of aging, robustly predicts age-related decline

**DOI:** 10.1101/2025.09.19.25336152

**Authors:** Swier Garst, Lieke Kuiper, Erik van den Akker, Niels van den Berg, Mohsen Ghanbari, Simon Mooijaart, Marian Beekman, Marcel Reinders, Eline Slagboom, Joyce van Meurs

## Abstract

Chronological age overlooks the heterogeneity in aging. In response, a wide range of molecular aging biomarkers has been developed to better capture an individual”s aging rate. Yet, a comprehensive comparison of modeling choices in the development of these biomarkers is lacking. In this study, we trained aging biomarkers on the Rockwood frailty index (FI) and all-cause mortality using UK Biobank Olink proteomics and metabolomics (^1^H-NMR) data (*n*=40,696). We systematically established the impact of model choice, target outcome, and molecular data source on several age-related outcomes. From this, we developed ProteinFrailty (ProtFI), an elastic net model using a minimal set of proteins to predict FI. ProtFI outperformed established aging biomarkers in relation to diverse outcomes, including incident cardiovascular disease, handgrip strength, and self-rated health, both in internal validation and two Dutch external cohorts (*n*=995, *n*=500). Our findings show that an efficient frailty-trained proteomic biomarker robustly predicts age-related decline.

## 1 Introduction

As individuals age, their risk of functional decline and disease increases^1,2^. However, aging is a heterogeneous process: individuals of the same chronological age may age at different rates and in different ways, resulting in varying risks for physical, cognitive, and functional outcomes^3^. Chronological age is commonly used as a proxy for aging but cannot, by definition, account for individual variation in age-related risk. As a result, the concept of biological age was introduced, aiming to reflect the pace and trajectory of aging on an individual level^4,5^.

Alternatively, the concept of frailty describes a state of increased vulnerability to stressors, resulting from decline across multiple physiological systems^6^. Among these, the Rockwood frailty index, further referred to as the frailty index, is one of the most widely used frailty measures^7^. Although distinct in concept, quantified frailty scores are frequently used in research and clinical practice as practical proxies for biological age^5^.

In addition to clinical assessments such as frailty, molecular or *omics* biomarkers have more recently been developed to reflect different aspects of biological aging^4,8^. Aging biomarkers are often derived from blood measurements and can span various molecular layers, each reflecting different aspects of the aging process^4^. Since the conception of the Horvath Clock in 2013^9^, numerous epigenetic clocks have been introduced, some of which incorporate clinical markers to enhance predictive accuracy^10^. In parallel, aging biomarkers derived from other omics data − including metabolomics^11^, transcriptomics^12^, proteomics^13^, and microbiome profiles^14^ − have emerged as alternative approaches to assess biological aging. Among these, transcriptome and epigenome signatures derived from blood cells are particularly indicative of cellular states that reflect systemic physiological conditions^10,12^. Proteins and metabolite signatures, on the other hand, more directly reflect the functional state of peripheral organs^15–17^.

First-generation *omics*-based aging biomarkers, known as “aging clocks”, were designed to estimate chronological age. In contrast, second-generation aging biomarkers are trained on health-related metrics, such as composite scores of disease burden or mortality risk^11,18^. Studies have demonstrated that biomarkers trained on mortality and age-related health data outperform those solely trained on chronological age in predicting health outcomes^19,20^. Given the broad spectrum of physiological impairments captured by the frailty index, we hypothesize that a proteomics-based aging biomarker trained to predict frailty could provide a robust and mechanistically relevant tool for assessing biological aging. Previous studies have attempted a similar approach, but either lack analysis regarding associations beyond frailty itself^21^, or are missing external validation^21,22^.

More recently, advances in machine learning have transformed aging biomarker development by enabling more complex, multi-variate approaches. Despite methodological advancements in aging biomarker development, practical implementations of aging biomarkers vary widely across the field. More critically, systematic work exploring key steps in biomarker development, such as model choice and training target selection, is lacking. Therefore, such choices in existing biomarker work remain mostly underscrutinized.

Furthermore, challenges remain in validation and standardization. Many new biomarkers lack rigorous external replication, and are rarely benchmarked against established biomarkers, making it difficult to assess their relative effectiveness. Consequently, a growing array of biomarkers based on single or multi *omic-*data has emerged^4,8^, but without systematic comparisons, their clinical relevance and reliability remain uncertain.

In this study, we developed aging biomarkers based on using Olink proteomics and Nightingale Health (^1^H-NMR) metabolomics data. We assessed the impact of model choice, target outcome, and molecular data source, identifying key factors that influence biomarker performance. Based on this analysis, we introduce two novel aging biomarkers: ProteinFrailty (ProtFI) and ProteinMortality (ProtMort), which leverage a minimal subset of proteins to predict frailty and mortality, respectively. By comparing ProtFI and ProtMort to previously established aging biomarkers, we demonstrate that an efficient protein-based approach, trained to predict frailty, provides robust and generalizable insights into aging phenotypes. By validating both aging biomarkers in two independent cohorts, we underscore the need to establish biomarker robustness across independent cohorts and populations

## 2 Results

### 2.1 Study Outline

We systematically investigated the use of proteomics for developing aging biomarkers. We evaluated key design choices in this process, structured into three main components: 1) model selection, 2) protein selection, and 3) prediction target. To assess model selection, we constructed aging biomarkers using ElasticNet (EN) models and deep Feedforward Neural Networks (FNN). These models were selected because they represent popular choices within two categories of models, being simple but linear (EN), or complex and non-linear (FNN). These models were trained on various subsets of plasma proteins from the UK Biobank (UKB)^23,24^, ranging from four Olink panels (1428 proteins), to the cardiometabolic panel only (344 proteins), to a subset of the cardiometabolic panel (20/27 proteins). Each biomarker was designed to predict either the frailty index (determined at the same time point as the protein measurement) or allcause mortality. In the UK Biobank, the frailty index consisted of 49 categorical deficits, which are categorized into sensory, cranial, mental well-being, infirmity, cardiometabolic, respiratory, musculoskeletal, immunological, cancer, pain and gastrointestinal related deficits^25,26^ (specific frailty index deficits used in this study are detailed in Methods and Supplementary Tables 1 and 2).

All aging biomarkers were trained, tested, and internally and independently validated on the UKB data, and externally validated in the Rotterdam Study (RS)^27^ and the Leiden Longevity Study (LLS)^28^. Table 1 summarizes the population characteristics. The UKB actively recruited National Health Service (NHS) patients^23^, and exhibits a strong healthy volunteer bias^29^. In contrast, the RS is a population-based study^27^, while the LLS comprises offspring of long-lived individuals and their partners^28^. For further details, see the study population section in the Methods. Fig. 1 shows a graphical representation of the study outline. For a selected set of aging biomarkers, we conducted association analysis with a range of age-related health outcomes, chosen based on their availability with statistical power across the three cohorts. All reported effect estimates in the UKB refer to those obtained from the internal validation set.

**Table 1:** Population characteristics. Values represent mean and standard deviation between parentheses for continuous variables (chronological age, frailty index, BMI, maximum handgrip strength, percentage of lymphocytes, and percentage of monocytes), median and the interquartile range between brackets for both the mortality as well as the incident cardiovascular disease follow-up time, and the number of participants and the percentage between brackets for the categorical variables (number of women, died during follow-up, highest attained eduction, current smoker, prevalent cardiovascular disease, incident cardiovascular disease, being part of the multi-omics set, and being non-randomly selected by the UK Biobank Pharma Proteomics Project (PPP) as participant).

|  | UK Biobank | Rotterdam Study | Leiden Longevity Study |
| --- | --- | --- | --- |
| Number of participants | 40,696 | 995 | 500 |
| Age in years | 57.5 (8.2) | 62.7 (5.8) | 66.0 (6.7) |
| Number of women | 21,941 (53.9) | 568 (57.1) | 252 (50.4) |
| Frailty index (0-1) | 0.13 (0.07) | 0.13 (0.07) | Not available |
| Died during follow-up | 4,497 (11.1) | 67 (6.7) | 95 (19.0) |
| Years of follow-up for all-cause mortality | 13.7 [13.0, 13.5] | 10.3 [9.6, 10.7] | 13.8 [13.4, 14.1] |
| Highest attained education |  |  |  |
| *Primary education* | 7,293 (17.9) | 68 (6.8) | 17 (3.4) |
| *Lower vocational or intermediate general education* | 11,079 (27.2) | 323 (32.5) | 119 (23.8) |
| *Intermediate vocational or secondary education* | 8,844 (21.7) | 280 (28.1) | 190 (38.0) |
| *Higher vocational education or university* | 12,764 (31.4) | 321 (32.3) | 168 (33.6) |
| BMI in kg/m<sup>2</sup> | 27.5 (4.8) | 27.3 (4.4) | 25.5 (3.7) |
| Current smoker | 4,357 (10.7) | 133 (13.4) | 31 (6.2) |
| Maximum handgrip strength in kilogram | 32.5 (11.3) | 31.2 (10.3) | 40.3 (11.7) |
| Self-rated health poor | 11,058 (27.2) | 85 (8.5) | 49 (9.8) |
| High blood pressure | 11,563 (28.4) | 570 (57.3) | 119 (23.8) |
| Prevalent cardiovascular disease | 2,040 (5.0) | 83 (8.3) | 31 (6.2) |
| Incident cardiovascular disease | 3,587 (8.8) | 60 (6.6) | 28 (7.3) |
| Years of follow-up for incident cardiovascular disease | 11.7 [11.0, 12.5] | 6.5 [6.0, 6.9] | 5.0 [4.4, 5.9] |
| Percentage of lymphocytes | 28.8 (7.7) | 35.0 (7.8) | Not available |
| Percentage of monocytes | 7.1 (2.6) | 6.4 (2.0) | Not available |
| Part of multi-omics set | 21,434 (52.7) | 583 (58.6) | 500 (100) |
| Selected as PPP participant | 4,898 (12.0) | Not applicable | Not applicable |

**Fig. 1:**
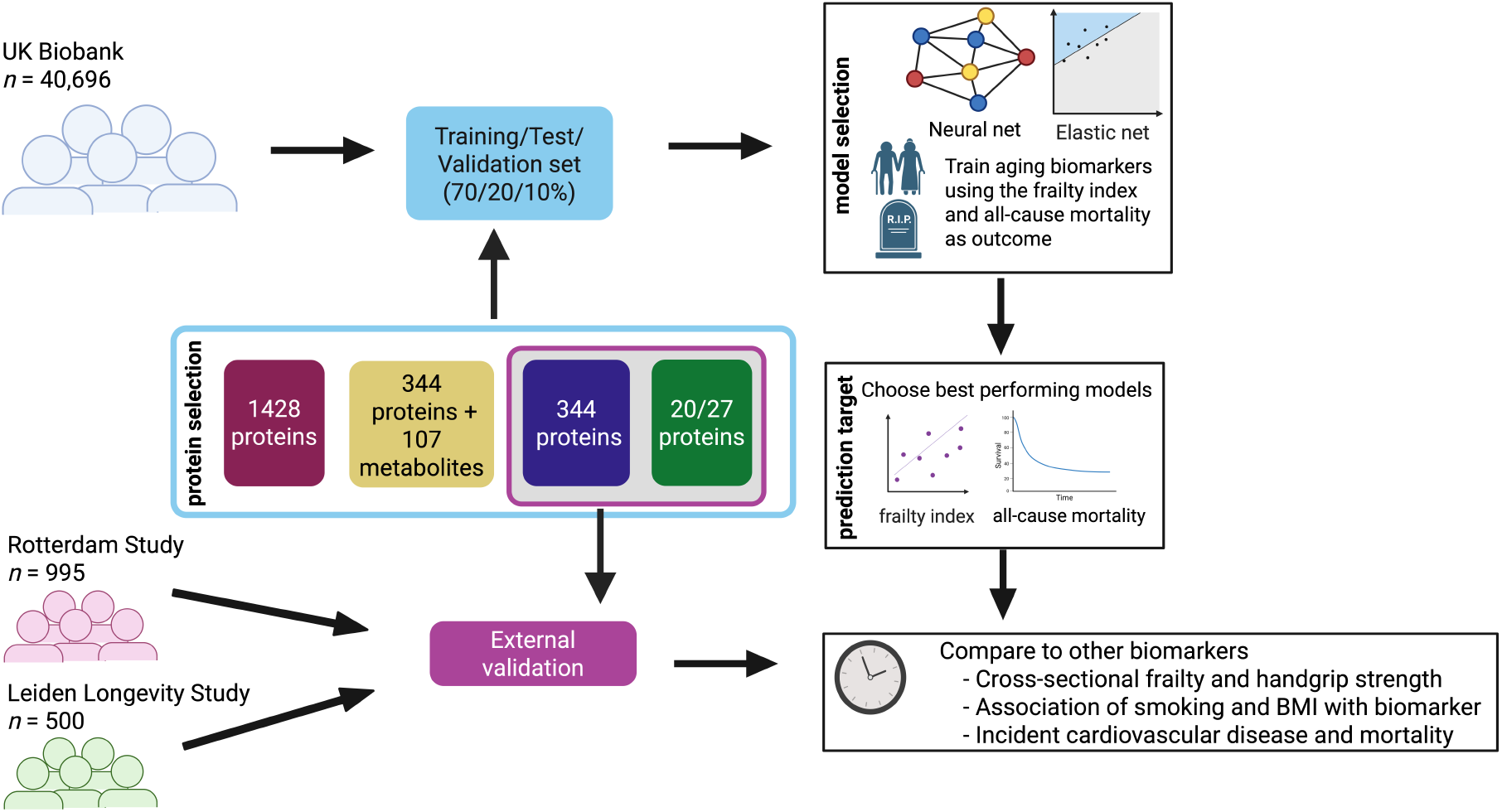
Overview of the study design. We used proteomics data from the UK Biobank as training data for all biomarkers. By creating a 70/20/10 training/test/validation split, we investigated the effect of the number of proteins included in a biomarker, as well as the effect of including metabolomics data. Furthermore, we compared linear ElasticNet and non-linear Feedforward Neural Network models to investigate the effect of using more complex models for biomarker training. We trained all our models to either predict the frailty index or mortality, in order to investigate the effect of training outcome. Finally, biomarkers based on the two smallest protein subsets were externally validated in two cohorts, the Rotterdam Study and the Leiden Longevity Study (as well as the validation set in the UK Biobank), by means of association analyses on several aging-related phenotypes.

### 2.2 Comparison between linear and deep learning models shows no benefit for more complex models

We first evaluated the impact of model complexity on aging biomarker performance. We started by using the set of four Olink panels, together making up 1428 proteins (allprot), and trained aging biomarkers based on outcomes and modelling approach. Markers were trained to either predict all-cause mortality or the frailty index, using two approaches: a linear ElasticNet (EN) model and a Feedforward Neural Network (FNN). This resulted in four biomarkers: EN trained on the frailty index and mortality (EN-allprot-FI and EN-allprot-mort), and FNN trained on the frailty index and mortality (FNN-allprot-FI and FNN-allprot-mort).

To assess model performance, we employed bootstrapping with replacement, generating 100 resampled training sets. For each resampled set, we applied the predetermined hyperparameters and evaluated performance metrics on the independent validation set, including *R*^2^ score for the frailty index as outcome and the Concordance-index (C-index) for mortality as outcome. We then compared the distribution of these performance metrics between the resulting EN and FNN aging biomarkers. Bootstrapping revealed a significant difference in performance (p-value after false discovery rate adjustment (pFDR) < 0.001), favoring the EN model over the FNN model for both the frailty index and mortality (Fig. 2). On average, the EN biomarkers exhibited an *R*^2^ that was 0.034 (standard deviation (SD) 0.008) higher and a C-index that was 0.015 (SD 0.010) higher than their FNN counterparts. These findings suggest that increased model complexity, as in FNNs, does not enhance performance.

**Fig. 2:**
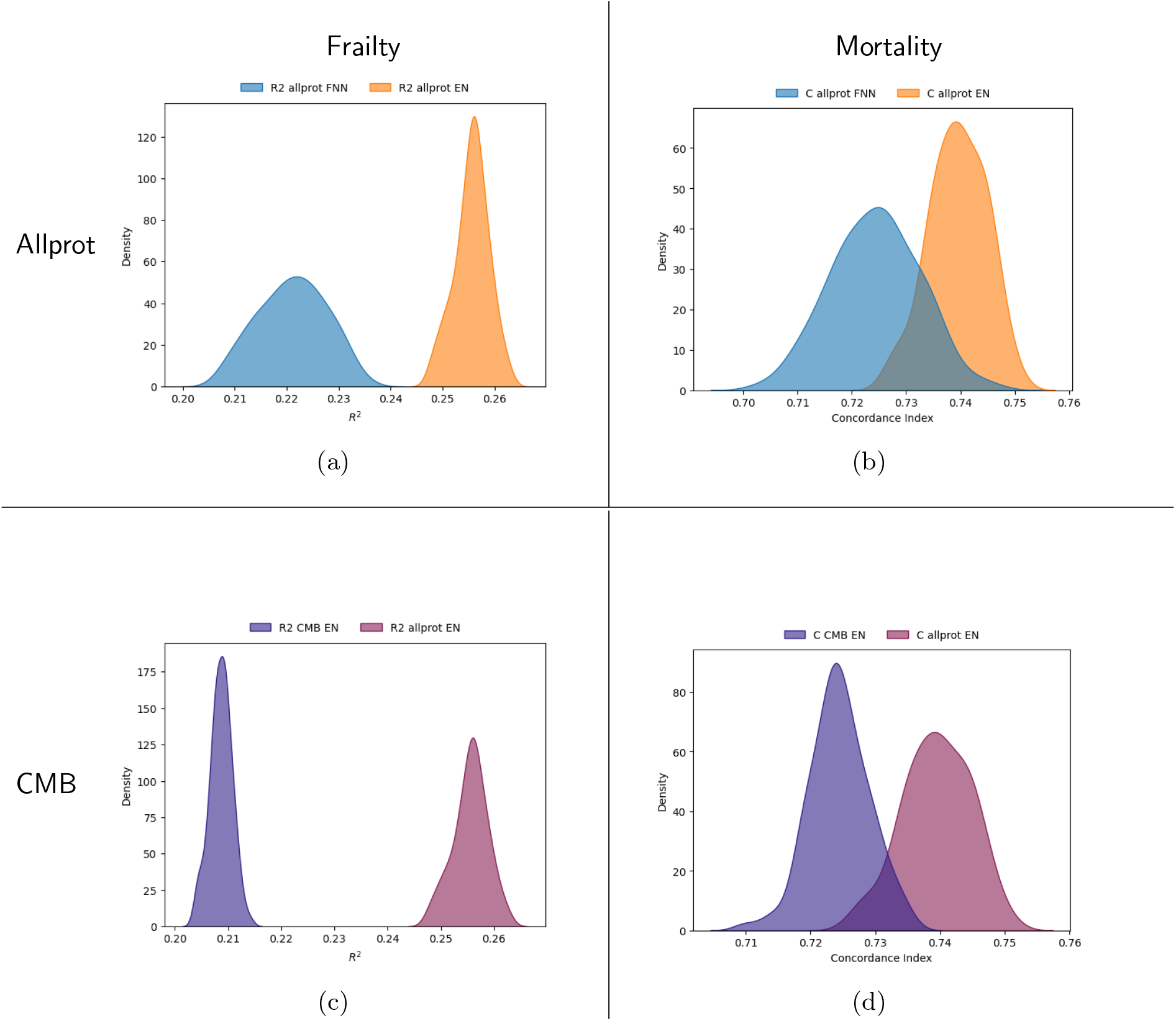
Density plot showing (a) & (c) *R*^2^ score (when trained on frailty) and (b) & (d) C-index (when trained on mortality) for aging biomarkers, comparing ElasticNet (EN, orange) and Feedforward Neural Network (FNN, blue) in (a) & (b), and the allprot (purple) and cardiometabolic (CMB, red) protein sets when training EN in (c) & (d). Distributions reflect performance on the fixed 10% independent validation set across models trained with the pre-selected hyperparameters on 100 bootstrap resamples of the combined training and test set (90% of the original UKB data on which the models were optimized).

### 2.3 Associations with non-training endpoints remain similar when reducing the number of proteins

For theoretical and future practical reasons, we asked the question how much redundancy exists in the information of the full set of proteins used so far. Therefore, we evaluated the impact of reducing the number of proteins used for aging biomarker training, decreasing the input from 1428 to the cardiometabolic (CMB) panel of 344 proteins (based on Olink panel distribution, see Methods for details). This selection was guided by the availability of this panel in the RS and LLS cohorts, enabling external validation. As in the previous analysis, we trained aging biomarkers using EN and FNN models on the UKB training set to predict either the frailty index or all-cause mortality, resulting in four CMB-based biomarkers: EN-CMB-FI, EN-CMB-mort, FNN-CMB-FI and FNN-CMB-mort. The performance metrics (*R*^2^ score or C-index) of the resulting CMB-based aging biomarkers on the independent UKB validation set were slightly, yet significantly, lower compared to the aging biomarkers trained on the full set of proteins (Fig. 2c and 2d, pFDR < 0.001). Specifically, the EN aging biomarker trained on all proteins (EN allprot) demonstrated, on average, a 0.047 (SD 0.003) higher *R*^2^ for the frailty index prediction and a 0.015 (SD 0.005) higher C-index for all-cause mortality prediction compared to the EN aging biomarker trained on the CMB panel (EN CMB).

In the UKB validation set, we then examined the associations per standard deviation (SD) increase in the age-accelerated biomarkers, which reflects the component of the biomarker independent of chronological age and indicates whether individuals are biologically older or younger than the population average, trained on either the allprot or CMB protein set with various age-related health indicators (see Methods, section 4.8 for details). These analyses were adjusted for sex, socio-economic status, smoking status, BMI, and cell counts. Figure 3 illustrates the associations between the aging biomarkers and their respective training targets: the frailty index and all-cause mortality. As expected, aging biomarkers generally exhibited stronger associations with the specific health indicator they were trained to predict—the frailty index for frailty index-trained biomarkers (top left, Fig. 3) and mortality for mortality-trained biomarkers (bottom right, Fig. 3). However, none of these differences were statistically significant after FDR correction (pFDR > 0.10).

**Fig. 3:**
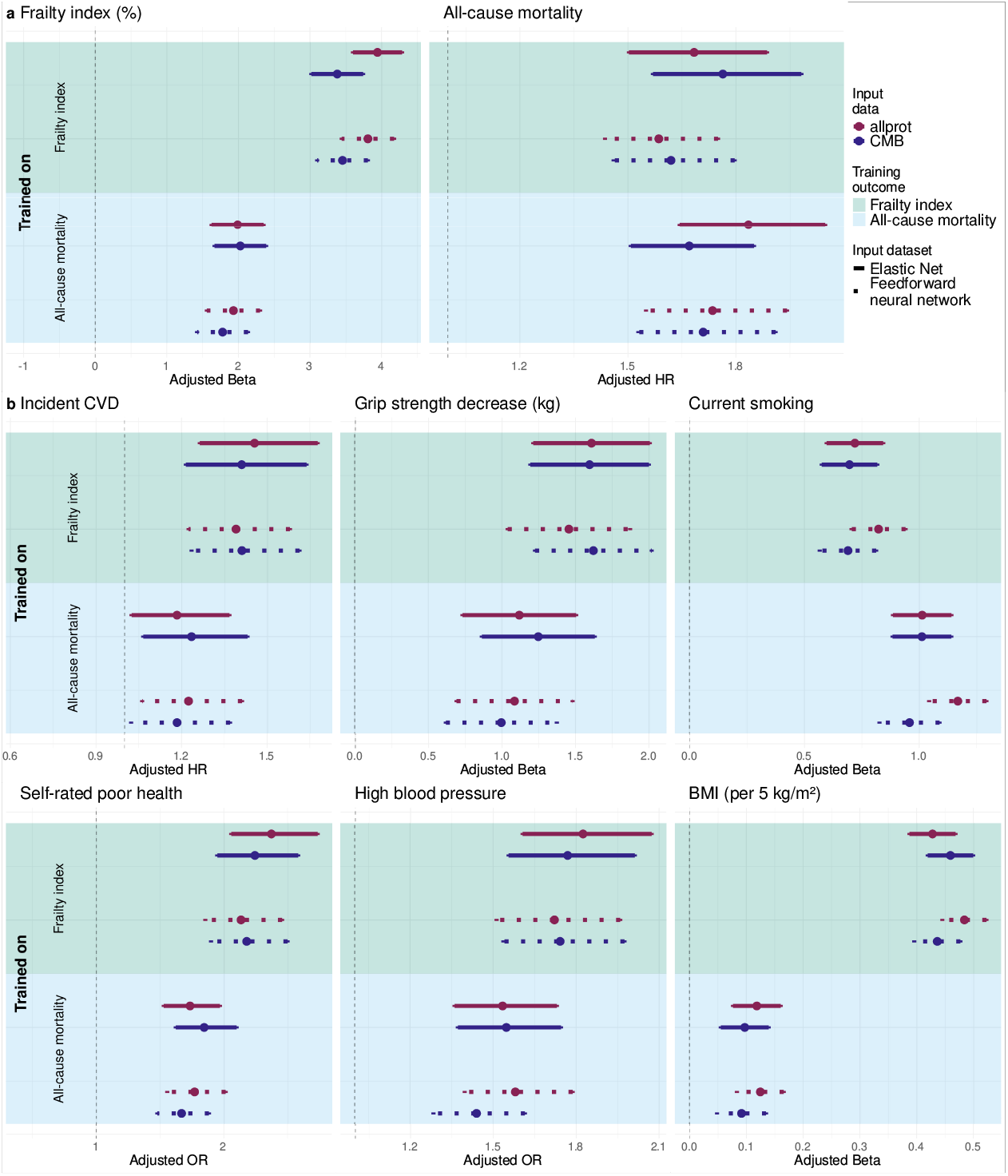
Health indicator associations of aging biomarkers trained on allprot and CMB proteins on the UK Biobank (UKB). The figure shows associations of protein-based aging biomarkers trained using four Olink panels (allprot, red) or only the cardiometabolic panel (CMB, blue) with: (a) the training endpoints, namely the frailty index and all-cause mortality, whereas (b) other age-related health indicators. Dots represent the point estimates. Solid and dotted error bars indicate 95% confidence intervals for EN and FNN biomarkers, respectively. The background color indicates the training outcome, with green (upper) for the frailty index and blue (lower) for all-cause mortality. All performance metrics were measured in the independent UKB validation set

For frailty index-trained biomarkers, the adjusted percentage increase in the frailty index per SD in the aging biomarker was 3.94 (95% confidence interval (CI): (3.60, 4.28)) for EN-allprot-FI and 3.38 (95% CI: (3.02, 3.74)) for EN-CMB-FI; the FNN-based aging biomarkers exhibited similar associations.

Similarly, mortality-trained biomarkers demonstrated slightly lower but comparable associations with mortality. The EN-allprot-mort biomarker was associated with a hazard ratio (HR) of 1.83 (95% CI: (1.64, 2.05)), while EN-CMB-mort had an HR of 1.67 (95% CI: (1.51, 1.85)). The FNN-based biomarkers followed a similar trend.

Notably, this pattern was not observed for the alternative training outcome (i.e., frailty when predicting mortality and vice versa; top right and bottom left, Fig. 3a, left panel), where differences between aging biomarkers trained on allprot or CMB were even smaller (pFDR > 0.99). When assessing associations with all-cause mortality, frailty index-trained biomarkers demonstrated equivalent performance when comparing the allprot and CMB protein sets. The EN-allprot-FI aging biomarker yielded a HR of 1.68 (95% CI: (1.50, 1.89)), while the EN-CMB-FI biomarker had an HR of 1.76 (95% CI: (1.57, 1.98)). The FNN-based aging biomarkers exhibited slightly lower HRs, though still comparable between the two protein sets.

The same held true in the reverse case (Fig. 3a, right panel), where mortalitytrained biomarkers were evaluated against the frailty index. Both EN- and FNN-based biomarkers showed similar performances across protein sets; the EN-allprot-mort biomarker showed an adjusted percentage increase of 1.99 (95% CI: (1.62, 2.35)), while the EN-CMB-mort biomarker had an increase of 2.03 (95% CI: (1.66, 2.39)). Altogether, this suggests that, although using more proteins increases performance on the training target, it does not benefit associations with other age-related health indicators.

Subsequently, we evaluated the impact of data source on biomarker performance. We investigated the integration of Olink proteomics data (CMB panel only) with 107 non-derived Nightingale Health (^1^H-NMR) metabolite markers using three approaches: (i) direct concatenation of proteins and metabolites, (ii) applying principal component analysis (PCA) separately to each data type followed by concatenation of the first ten or twenty principal components of each modality, and (iii) AJIVE, which finds a common subspace between the two data modalities (methods detailed in Supplement 5.2). However, none of the integration methods, whether using FNN or EN, resulted in higher aging biomarkers” performance metrics (*R*^2^ or C-index); see Supplement 5.2.

### 2.4 Associations with other health outcomes

Next, we investigated to what extend the frailty index or mortality trained markers predicted relevant age-related health indicators. Fig. 3b demonstrates that a model trained on the frailty index predicts these health outcomes better than a model trained on mortality, with the exception of smoking. Secondly, regardless of the training method, biomarkers derived from the reduced set of proteins yield comparable associations (pFDR > 0.14).

Furthermore, for some health indicators, such as the association between frailty index-trained FNN biomarkers and self-rated poor health, the FNN-CMB biomarker showed slightly higher point estimates (odds ratio (*(OR)*) = 2.13, 95% CI: (1.85, 2.45) for FNN-allprot-FI vs. *OR* = 2.18, 95% CI: (1.90, 2.50) for FNN-CMB-FI). Similarly, for incident cardiovascular disease, the FNN-CMB-FI biomarker yielded higher point estimates. This indicates that this subset of proteins is able to retain important information with respect to broader aging factors.

### 2.5 Forward Feature Selection is an effective method in selecting highly associative proteins for an aging biomarker

Biological age markers ideally capture a range of aging-related health outcomes to indicate overall health of individuals with rather a minimum than a maximum of molecular features. We observed that the subset of CMB proteins as compared to allprot can retain equivalent associations with a range of health outcomes beyond those they were trained on. Therefore, we aimed to identify a “minimal selection” of proteins that maintain robust associations with various age-related health indicators tested earlier. To achieve this, we applied a Forward Feature Selection (FFS) approach to identify proteins from the CMB protein set that incrementally increase goodness-of-fit metrics by more than 0.001, i.e. *R*^2^ in the case of the frailty index (27 proteins plus chronological age) or the C-index in the case of all-cause mortality (20 proteins) in the UKB training set (see Methods for details). The final protein lists are available in Supplementary Table 4. Using these minimal protein sets, we created aging biomarkers using ElasticNet, named ProtFI and ProtMort, predicting the frailty index and all-cause mortality, respectively. For comparative reasons, we also trained Feedforward Neural Network models on the same reduced protein sets to predict frailty (FNN-FFS-FI) and mortality (FNN-FFS-mort). We assessed the associations of the resulting FFS aging biomarkers with the frailty index, all-cause mortality, and various health indicators in the independent UKB validation set.

Reducing the number of proteins from 344 to 27 for frailty index-based models, and to 20 for all-cause mortality models, resulted in some general attenuation of associations with the respective training outcomes (Fig. 4a). However, these differences were not statistically significant for any aging biomarker (all pFDR > 0.86). When restricting the analysis to 27 proteins and age, the adjusted effect size (*β*) on the frailty index itself for the frailty index-trained EN biomarker decreased from 3.38 (95% CI: (3.02, 3.74)) for EN-CMB-FI to 3.02 (95% CI: (2.64, 3.39)) for ProtFI. A similar attenuation was observed for the FNN-based aging biomarkers.

**Fig. 4:**
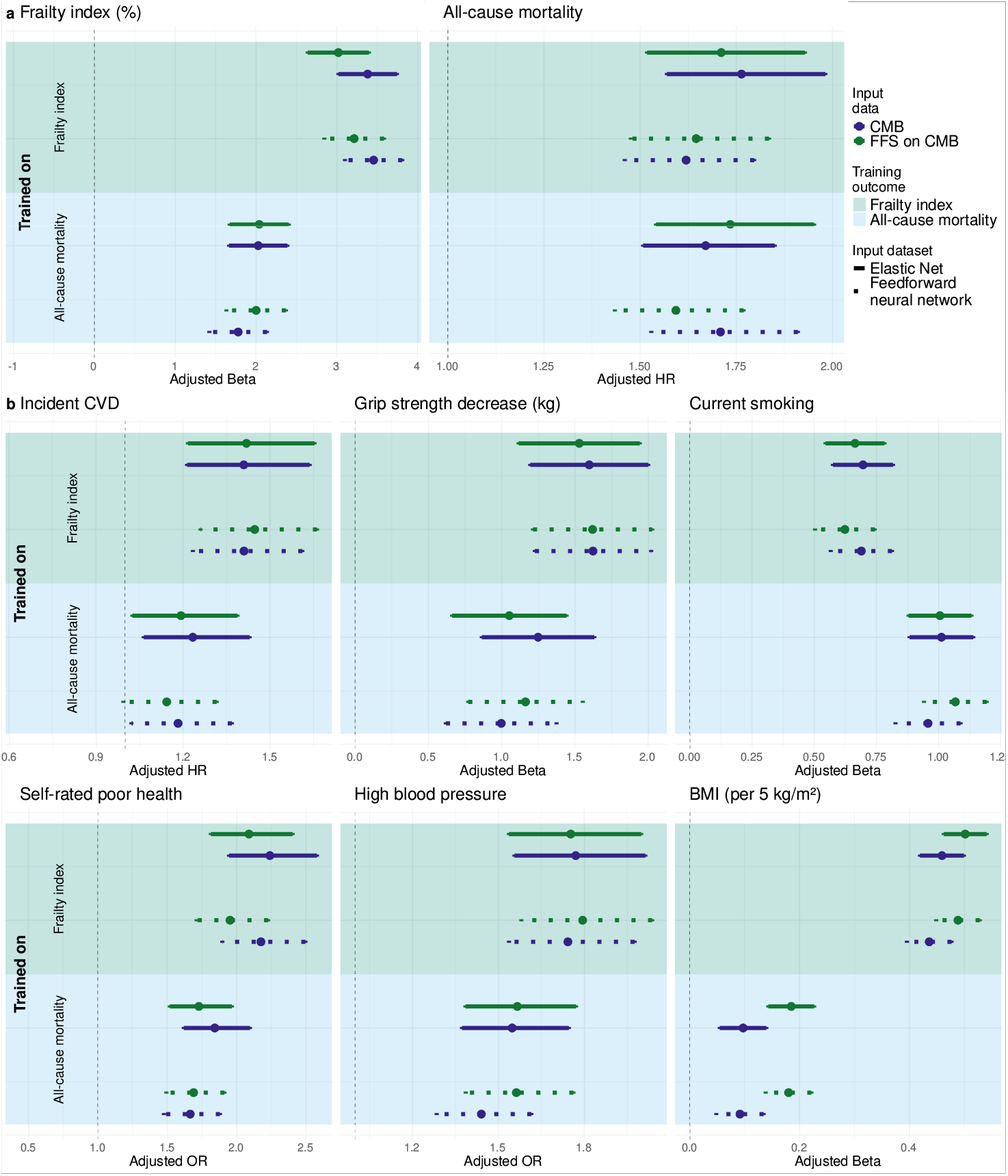
Health indicator associations of aging biomarkers trained on CMB and FFS-selected proteins. The figure shows associations of protein-based aging biomarkers trained using the cardiometabolic (CMB) panel or the forward feature selected CMB proteins (FFS) with: (a) the training endpoints, namely the frailty index and all-cause mortality, and (b) other age-related health indicators. See Fig. 3 for figure elaboration. All performance metrics were measured in the independent UKB validation set

For all-cause mortality trained biomarkers, the HR for mortality prediction for ProtMort was slightly higher than for the EN-CMB-mort estimate (ProtMort: *HR* = 1.73, 95% CI: (1.54, 1.95) vs. EN-CMB-mort: *HR* = 1.67, 95% CI: (1.51, 1.85)), though this difference was not statistically significant. In contrast, for mortality-trained FNN biomarkers, a greater number of proteins resulted in a stronger association with the training outcome. This suggests that, overall, using a larger protein set enhances predictive performance for the training outcome, consistent with findings presented in Fig. 3.

Fig. 4b further compares the associations of FFS and CMB aging biomarkers with various health outcomes. The full comparison between allprot, CMB and FFS based biomarkers can be found in Supplementary Fig. 1. Despite being trained on fewer than one tenth of the proteins used in the CMB aging biomarkers, both EN- and FNN-trained FFS aging biomarkers showed no significant attenuation in their associations with health outcomes compared to CMB-trained biomarkers within the same modeling framework and training outcome (all pFDR > 0.71). In some cases, the point estimates for the FFS aging biomarkers were even higher than those for the CMB aging biomarkers, such as for the association with incident cardiovascular disease in frailty index-trained aging biomarkers (*HR* = 1.45, 95% CI: (1.26, 1.66) for FNN-FFS-FI, vs. *HR* = 1.41, 95% CI: (1.23, 1.61) for FNN-CMB-FI) and prevalent high blood pressure in mortality-trained aging biomarkers (*OR* = 1.56, 95% CI: (1.39, 1.76) for FNN-FFS-mort vs. *OR* = 1.44, 95% CI: (1.28, 1.62) for FNN-CMB-mort). These findings further suggest that protein-based aging biomarkers trained on a reduced protein set maintain their ability to capture key aspects of the aging process, despite the substantial reduction in the number of proteins.

### 2.6 Comparison with published aging biomarkers

Given that biomarkers developed using less complex models with a reduced number of proteins demonstrated robust associations with aging phenotypes, we proceeded with the EN-trained FFS aging biomarkers, ProtFI (frailty index-trained) and ProtMort (mortality-trained). We then compared ProtFI and ProtMort with previously published aging biomarkers (see Methods for details). Specifically, we benchmarked ProtFI and ProtMort against the proteomics-based aging biomarker ProteinAge and ProteinAge20, trained on chronological age^30^, and two mortality-trained biomarkers, the ProteinMortality Score^31^, and the Proteomic Aging Clock (PAC)^32^. Additionally, we included two metabolomics-based aging biomarkers, both of which predict mortality: The Metabo Mortality Score^31^ and MetaboHealth^11^, which − unlike the other biomarkers was not − trained in the UK Biobank. Since previous studies identified GDF15 as a key protein associated with mortality^17^, and given that it was the most important protein selected by our FFS procedures for both the frailty index and all-cause mortality, we also evaluated GDF15 alone.

Fig. 5 demonstrates that, despite using fewer proteins than PAC (128 proteins, derived from 2923 proteins), the ProteinMortality Score (201 proteins) or ProteinAge (204 proteins), ProtFI exhibited the strongest associations with the frailty index, incident cardiovascular disease, self-rated poor health, prevalent high blood pressure, and was most prominently associated with BMI. The differences in association between ProtFI and the next most associative model (per endpoint) remain significant for the frailty index and the association with BMI. While stronger associations for ProtFI might be expected for indicators included in the frailty index (e.g., high blood pressure and self-rated poor health), similar trends were observed for independent measures like incident cardiovascular disease and handgrip strength.

**Fig. 5:**
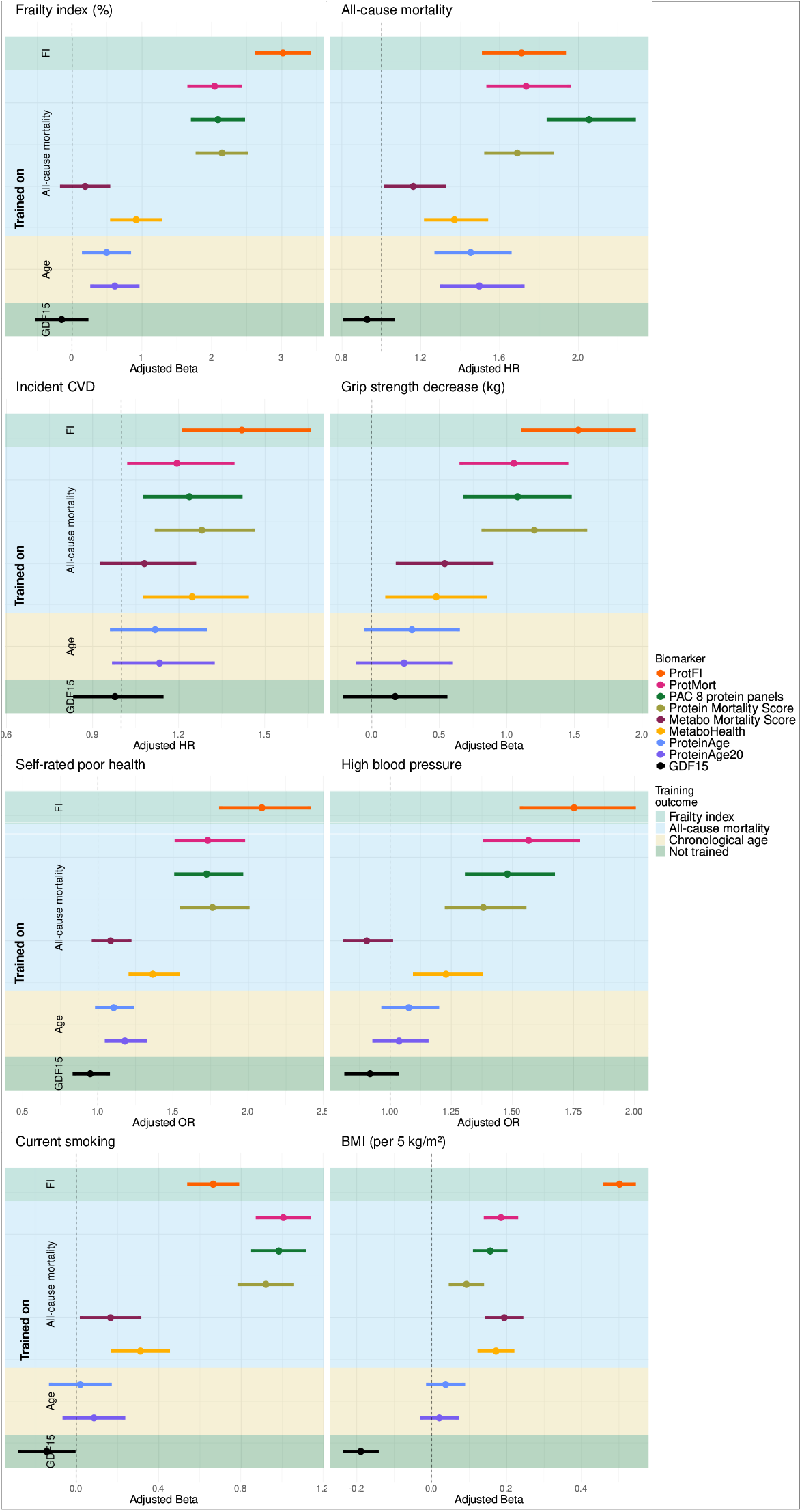
Comparison of associations of ProtFI and ProtMort with previously published aging biomarkers. Dots represent the point estimates. Error bars are displayed to show the 95%-confidence interval. The background color indicates the training outcome, with green (upper) for the frailty index and blue (second) for all-cause mortality, yellow (third) chronological age, and dark green (bottom) GDF15, a non-trained agsing biomarker. Training outcome is also displayed on the y-axis. Adjusted beta and OR are estimated in the independent UKB validation set.

In contrast, for all-cause mortality, PAC showed the strongest association (ProtFI: *HR* = 1.71, 95% CI: (1.52, 1.93) vs. PAC: *HR* = 2.05, 95% CI: (1.85, 2.28)). Furthermore, smoking was strongest associated with mortality-trained proteomics-based biomarkers, with ProtMort showing the highest effect size (ProtMort: *β* = 1.00, 95% CI: (0.88, 1.13) vs. Protein Mortality Score *β* = 0.92, 95% CI: (0.79, 1.05); PAC: *β* = 0.98, 95% CI: (0.86, 1.05)).

### 2.7 Validation in external cohorts shows generalizability of biomarkers

We validated our aging biomarkers in two independent external cohorts, the Rotterdam Study (RS) and the Leiden Longevity Study (LLS). Given that the CMB panel was the only proteomic dataset available in these cohorts, our validation efforts focus on aging biomarkers trained on this subset of proteins, as well as ProtFI and ProtMort. External validation in these cohorts demonstrated that biomarkers trained using FFS-selected proteins exhibited effect sizes comparable to those obtained using the full CMB panel (all pFDR > 0.60), as shown in Supplementary Fig. 2 and Supplementary Table 5.

Consistent with our approach in the UKB, we also compared ProtFI and ProtMort with the metabolomics-based aging biomarker MetaboHealth. Additionally, in a subset of RS, we extended our comparisons to epigenetic biomarkers trained on chronological age^9,33^, or mortality-associated phenotypes^18^, providing a broader context for evaluating their performance relative to well-established aging biomarkers.

Fig. 6 (Supplementary Table 5) presents the associations of the aging biomarkers in both the multi-omics subset of RS and LLS, while results from the full RS cohort can be found in Supplementary Table 6. Findings in these external validation cohorts were consistent with those in the UKB, with ProtFI demonstrating stronger associations with aging-related phenotypes compared to age-trained epigenetic biomarkers, including DNAm Horvath and DNAm Hannum. In particular, ProtFI exhibited the strongest associations with aging phenotypes regardless of whether they were self-reported or objectively measured. The only exception was the association with current smoking, which was consistent with our findings in the UKB. These results highlight the generalizability of ProtFI and ProtMort to an independent cohort and across diverse populations, reinforcing their ability to robustly reflect and predict the aging process.

**Fig. 6:**
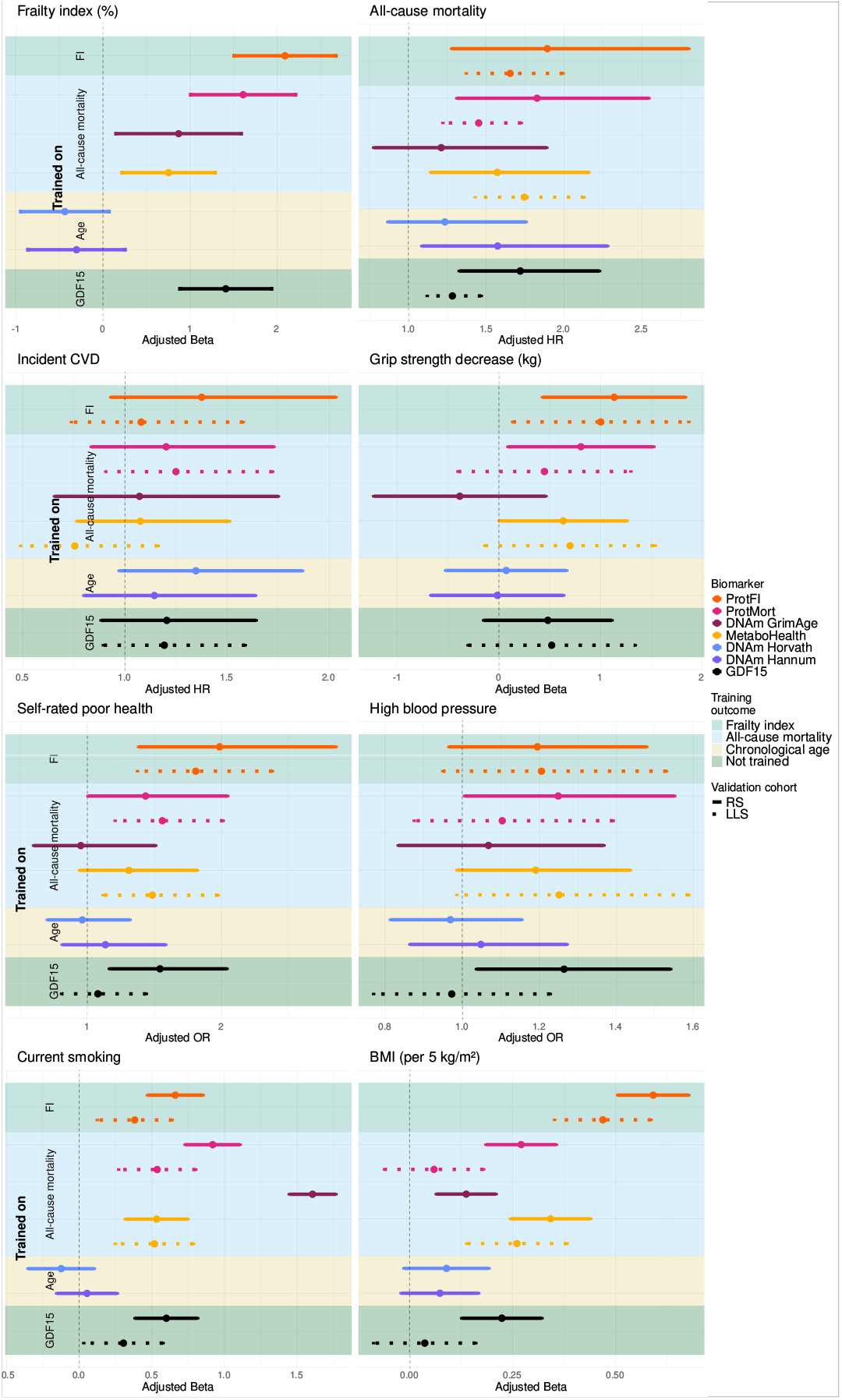
External validation of aging biomarkers in LLS and RS with different health outcomes. The figure shows associations of different aging biomarkers. Dots represent the point estimates. Solid and dotted error bars indicate 95% confidence intervals for the Rotterdam Study (RS) and Leiden Longevity Study (LLS), respectively. The background color indicates the training outcome, with green (upper) for the frailty index and blue (second) for all-cause mortality, only in RS, yellow (third for RS) chronological age, and dark green (bottom) GDF15, a non-trained aging biomarker. Training outcome is also displayed on the y-axis.

## 3 Discussion

This study systematically compared aging biomarker development with respect to model selection, training outcome, and input data. As a result, we present ProtFI, a proteomics-based aging biomarker trained to predict the frailty index, showing promising associations with various health indicators.

We did not observe clear advantages of neural networks over simpler linear models. Furthermore, when training on either the frailty index or all-cause mortality and evaluating associations with the same outcome, biomarkers performed best when including as many proteins as possible. When assessing associations with outcomes other than the training outcome (e.g., handgrip strength or self-rated poor health), however, biomarkers trained on the frailty index outperformed those trained on mortality, suggesting that the frailty index may provide a more comprehensive representation of biological aging. Moreover, simpler biomarkers based on a subset of cardiometabolic proteins or a small number of proteins selected through forward feature selection showed broadly similar associations with health outcomes beyond the training outcomes compared to aging biomarkers trained on the full proteomics set. In fact, when predicting outcomes different from the one the biomarkers were trained on, such as using frailty-trained biomarkers to predict mortality, those based on the full set of proteins performed slightly worse than the cardiometabolic ones, suggesting that the full proteomics set biomarkers may be somewhat overtrained or capture less relevant features. We introduced two biomarkers, ProtFI and ProtMort, which are based on a minimal set of proteins − 27 for ProtFI and 20 for ProtMort − from the forward feature selected protein set to predict the frailty index and mortality, respectively. ProtFI outperformed established aging biomarkers in association with aging phenotypes. Finally, both CMB- and FFS-based biomarkers were validated in independent cohorts, supporting the robustness and generalizability of their associations across populations.

Our findings reveal that within this dataset, the use of more complex models, such as neural networks, did not improve predictive performance for age-related clinical phenotypes in broad. This result is consistent with existing literature suggesting that neural networks struggle with tabular data, where traditional statistical models or simpler machine learning approaches tend to perform more effective^34^. While deep learning for tabular data remains an active area of research, with recent advances exploring the use of foundation models for such data^35^, further developments are needed to assess its broader applicability in this context.

Training biomarkers on the frailty index resulted in superior predictive performance for most endpoints, even for those not directly related to the frailty index, such as handgrip strength and incident cardiovascular disease. This is most likely due to the broader and more nuanced nature of frailty as a training target compared to mortality. We specifically chose the frailty index as a training outcome because it can detect differences even among relatively healthy individuals, in contrast to more physical frailty measures such as the frailty phenotype, which categorizes individuals as frail or not frail per component^36–39^. However, we did not compare predictive performance on various health outcomes among biomarkers trained on different frailty measures, which would be an important direction for future research. For outcomes with a strong association to mortality, such as smoking^40,41^, biomarkers trained directly on mortality outperformed those trained on frailty. This finding underscores the importance of determining the objective of developing an aging biomarker in advance, as the choice of training target should align with the biomarker”s intended function. It”s important to note that differences in statistical power between mortality and frailty index-based models may also influence these results, as the number of mortality cases impacts the training power of mortality, and the high degree of censoring in the training data could affect model training and evaluation.

Besides the selection of training target, our results emphasize the significance of considering the intended use of a biomarker when choosing the number of proteins to include. In both our internal validation set as well as two independent validation cohorts, increasing the number of proteins generally improved performance for the specific endpoint the biomarker was trained on, such as mortality or frailty. However, this improvement did not always lead to better associations with other health outcomes. In some cases, the point estimates of the FFS biomarkers were even higher than those of CMB. This may be explained by the targeted selection of proteins in FFS that are more consistently associated with age-related decline, which may have improved the signal-to-noise ratio by excluding proteins with weaker or more diffuse associations. This highlights the point that a lower goodness-of-fit score (e.g., the concordance index or *R*^2^ value) does not necessarily indicate that the biomarker performs poorly for other health outcomes. While these metrics are useful for model comparison, they should not be the sole criterion for evaluating a biomarker”s relevance in risk assessment, particularly in population health management or clinical settings. Reducing the number of proteins, for example, may lower costs and facilitate broader implementation in the population, even if it slightly diminishes predictive accuracy in a population-based context. Future research should carefully balance the tradeoff between accuracy and practicality, considering the biomarker”s translational potential in real-world applications.

In our study, multi-omics biomarkers did not outperform single-omics biomarkers in terms of predictive accuracy. However, both MetaboHealth and proteinbased biomarkers demonstrated independent associations with outcomes, suggesting that integrating multiple omics layers may still offer valuable complementary insights. This inability to create multi-omics aging biomarkers that consistently outperform single-omic aging biomarkers developed on one molecular layer is consistent with previous efforts to create multi-omics aging biomarkers^31^. The added noise from integrating different molecular layers may dilute the additional information provided by these layers. Furthermore, our results align with previous work emphasizing that multi-omics approaches may enhance biological understanding rather than directly improving predictive performance^42,43^. As the field of multi-omics integration methods is rapidly evolving, its potential to improve aging biomarker performance may change in the future.

Several limitations of the current study warrant consideration. The UKB is relatively young and healthy, resulting in lower statistical power for mortality-based biomarkers and a less frail population for frailty index prediction. This raises questions about the generalizability of these results to diseased or older populations. However, the external validation in older cohorts (RS and LLS) demonstrates the robustness of the aging biomarkers developed in this study. With the exception of MetaboHealth, all aging biomarkers we compared were developed using data from the UKB. On the other hand, RS and LLS were included in the training of MetaboHealth, accounting for 13.7 % of its training set^11^. The stronger associations of MetaboHealth in LLS and RS compared to UKB, and the reverse trend for the other biomarkers we compared, suggest that the effect sizes observed for the other biomarkers may be inflated due to overlap with their training data. Additionally, while we incorporate Olink proteomic and ^1^H-NMR metabolomic information, we did not include other molecular layers to fully assess the robustness of our methodological findings across different molecular platforms.

Besides the cost reductions associated with using fewer proteins, selecting a minimal set of proteins may also enhance robustness. By focusing only on proteins strongly associated with the training target, we reduce the risk of overfitting to noisy or less relevant proteins, thereby improving the generalizability of the biomarker to external cohorts. Our external validation results in the RS and LLS cohorts support this hypothesis, further highlighting the benefits of using a smaller, carefully selected protein set.

In conclusion, this study presents a framework to systematically develop and compare aging biomarkers. Our approach underscores the utility of biomarkers that are designed to predict outcomes beyond those used in their training. We introduce robust aging biomarkers, ProtFI and ProtMort, which trained on only 27 and 20 proteins, demonstrate that using more proteins or more complex models does not necessarily lead to a better biomarker performance. Furthermore, our results highlight that in a rapidly expanding field, generalizability and external validation must take priority in aging biomarker development. This study provides a foundation for future research on aging biomarkers and potential applications in both population health and clinical settings.

## Acknowledgements

The authors want to thank M. Austin Argentieri for the extensive help with the ProteinAge biomarker. This research has been conducted using the UK Biobank Resource under Application Number 67864.

## Data Availability

Data from the UK Biobank can be accessed through a procedure described at https://www.ukbiobank.ac.uk/enable-your-research. The data from the Leiden Longevity Study and the Rotterdam Study are accessible for researchers upon request. Please find the required forms for the Leiden Longevity Study at: https://leidenlangleven.nl/data-access/. Requests for Rotterdam Study data should be directed towards the management team of the Rotterdam Study), which has a protocol for approving data requests. Because of restrictions based on privacy regulations and informed consent of the participants, data cannot be made freely available in a public repository.

## Code Availability

Python and R code for reproducing the results in this manuscript can be found at https://github.com/swiergarst/ProtFI

## 4 Methods

### 4.1 Study Population

#### UK BioBank

The UK Biobank is a prospective cohort study consisting of 500,000 participants recruited in the United Kingdom between 2006 and 2010. Invitations were sent to approximately 9.2 million individuals aged 40–69 years who lived within 25 miles (40 km) of one of 22 assessment centers across England, Wales, and Scotland, with 5.5% taking part in the baseline assessment^23^. We limited our UKB sample to participants with Olink Explore data available at baseline and who were randomly selected from the broader UKB population (n = 40,696). The data in the current study were accessed under application number 67864.

The UK Biobank received approval from the National Information Governance Board for Health and Social Care and the National Health Service North West Multicentre Research Ethics Committee.

#### Rotterdam Study

The Rotterdam Study (RS) is a prospective population-based cohort based in Ommoord, a suburb of Rotterdam, the Netherlands. The RS began in 1990 with 7,983 participants aged 55 and older and later expanded with additional cohorts: RS-II (in 2000, 3,011 participants aged 55 ≤), RS-III (in 2006, 3.932 participants aged 45 ≤), and RS-IV (in 2016, 3,005 participants aged 40 ≤). Participants underwent home interviews and standardized health assessments at baseline and follow-ups every 3 to 5 years, conducted by trained research staff in a dedicated research facility^27^. For this study, we measured the Olink Explore Cardiometabolic panel in RS-III participants who attended the research facility for the second visit, prioritizing those with the greatest overlap with metabolomics, gut microbiome, and DNA methylation data measured(n = 996), for which participants had previously been randomly selected. Despite this prioritization, not all selected participants had complete data across all molecular platforms.

The Rotterdam Study has been approved by the Medical Ethics Committee of the Erasmus MC (registration number MEC 02.1015) and by the Dutch Ministry of Health, Welfare and Sport (Population Screening Act WBO, license number 1071272-159521-PG). The Rotterdam Study Personal Registration Data collection is filed with the Erasmus MC Data Protection Officer under registration number EMC1712001. All participants provided written informed consent to participate in the study and to have their information obtained from their treating physicians.

#### Leiden Longevity Study

The Leiden Longevity Study (LLS), conducted between 2002 and 2006, includes 420 Dutch Caucasian families with at least two long-lived full siblings (≥ 89 years for men, ≥91 years for women) who were willing to participate. The study enrolled ≥ 991 siblings and cousins (first generation), as well as 1,671 of their children and 744 of their children”s partners (second generation)^28^. For this study, we included a subset of 500 second-generation participants and their partners who attended the third measurement of whom NMR-metabolomics was previously measured, and in whom the Olink Explore Cardiometabolic panel was assessed.

The Leiden Longevity Study protocol was approved by the Medical Ethical Committee of the Leiden University Medical Center before the start of the study (P09.140) at August 5^*th*^, 2009. The first participant was enrolled at August 24^*th*^, 2009. In accordance with the Declaration of Helsinki, the Leiden Longevity Study obtained informed consent from all participants prior to their entering the study.

### 4.2 Data preprocessing

#### Proteomic profiling

The Olink Explore assay (Olink, Uppsala, Sweden), based on the proximity extension assay technique, was used to quantify plasma levels of 2,923 proteins in the UK Biobank and 384 proteins in the Rotterdam Study and Leiden Longevity Study. The Olink Explore 3072 platform integrates eight Olink panels—Cardiometabolic, Cardiometabolic II, Inflammation, Inflammation II, Neurology, Neurology II, Oncology, and Oncology II. Based on data availability at the time of analysis and our overarching aim to maximize biomarker generalizability, we used the non-II panels for biomarker development (total of 1463 proteins before quality control). The II-panels were only included in the calculation of previously published aging biomarkers^30,32^ that incorporated proteins from these panels. In the Rotterdam Study and Leiden Longevity Study, only the Cardiometabolic panel was measured. Previous proteomic studies in the UK Biobank have described the profiling conducted in that cohort^24^.

In the Rotterdam Study and Leiden Longevity Study, proteomic analysis on EDTA plasma was performed by the Genomics Core Facility (www.genomicserasmusmc.nl) in Rotterdam following the manufacturer”s protocol. Data are presented as NPX (Normalized Protein eXpression) values, Olink”s relative protein quantification unit on a log2 scale. NPX values are calculated from matched counts using Next-Generation Sequencing (NGS) as the readout, providing a standardized measure of protein abundance. To enhance the comparability of NPX values across cohorts, all protein measurements were Z-transformed.

#### Quality Control

Quality control for the UKB proteomics data was done by UKB^24^. For the Rotterdam Study and Leiden Longevity Study proteomics data, the quality control followed the standard manufacturer”s procedure. In short, three internal controls are added to each sample, the Incubation control, the Extension Control, and the Amplification control. The Extension Control is used for the generation of the NPX values. The Incubation Control and the Amplification Control are used to monitor the quality of assay performance, as well as the quality of individual samples. Three external controls are included in each run, Plate Control (healthy pooled plasma), Sample Control (healthy pooled plasma), and Negative Control. The Plate Control is used for data normalization, the Sample Control is used to assess potential variation between runs and plates, and the Negative Control is used to calculate the Limit of Detection for each assay and to assess potential contamination of assays. To pass the quality control, a threshold of an average of 500 reads per combination of sample and protein was used. Additionally, for each protein per plate, the deviation of the median of the Negative Controls must be ≤ 5 standard deviations. One RS-participant failed quality control and was excluded from all analyses.

In the UK Biobank, there were after quality control by the cohort 52,704 participants with olink protein information available. We excluded participants with >2% missingness on their proteomic information, and participants with proteomic information from multiple visits. This resulted in 40,696 particiants whose proteomics data was used. Subsequently, we excluded proteins with >2% missing values, resulting in the removal of 36 proteins, leaving 1428 proteins for analysis. Further missing proteins were imputed using K-nearest-neighbor imputation, using the KNNImputer function of the SKLearn package, with k = 5.

The same procedure was followed in both external cohorts. In both RS and LLS, no participants were excluded due to missingness. However, three proteins that were not included in either ProtFI or ProtMort, namely BMP6, EPHX2, and PGLYRP1, failed the quality control in both studies. These proteins have been imputed by the proteins with the highest correlations to these proteins in the UK Biobank, respectively PDGFA, GSTA1, and RETN (see supplementary table 8 for a full list of highest correlating proteins in the UK Biobank).

#### Profiling other molecular layers

Metabolomic profiling of EDTA plasma was performed using the Nightingale platform. In the UK Biobank, the Nightingale 2020 assay was measured, whereas the Rotterdam Study and the Leiden Longevity Study used the 2016 assay, which was subsequently re-quantified to align with the Nightingale 2020 assay^44^. Additionally, DNA methylation patterns were measured in the Rotterdam Study, with further details provided elsewhere^20^.

#### Data splits

In the UK Biobank, we created a 70/20/10 split for all participants, representing the train/test/validation split (*n* = 28,487/8,138/4,071) for all proteinbased biomarkers. We also created a similar split for the subgroup of participants who had both proteomics and metabolomics information available (*n* = 15,060/4,321/2,053 for train/test/validation splits). We sampled the multi-omic set in such a way that each of the splits for the multi-omics dataset contains only participants of its respective split in the protein-only dataset (I.e. the 15,060 samples in the multi-omics training set are all also within the 28,487 samples of the proteomics-only training set); See Supplementary Figure 3 for details. We did not stratify on sex, as preliminary experiments (as well as earlier work)^30^ show no significant differences between sexes (Supplementary Table 7).

### 4.3 Ascertainment of covariates and outcomes

#### Ascertainment of primary outcomes

The Rockwood frailty index (FI), an accumulation measure of age-related deficits^26^, was calculated using the previously established methods^25,45^. The FI consisted of 49 deficits in the UK Biobank and 38 in the Rotterdam Study. The FI was calculated if a person had information on at least 20 deficits before multi-chain imputation as the number of deficits/total number of deficits in that cohort.

All-cause mortality was defined as participants who died from any cause during the total follow-up period, which concluded on the 30^*th*^ of November 2022 in the UK Biobank, the 5^*th*^ of July 2024 in the Rotterdam Study, and 8^*th*^ of April 2024 in the Leiden Longevity Study.

#### Ascertainment of covariates

Chronological age was calculated as the time between birth and blood sampling. In the UK Biobank, the exact date of birth was not available to researchers; therefore, we used the age at assessment as provided by the UK Biobank. Socioeconomic status was defined as the highest attained education following the UNESCO classification^46^. Current smoking status was defined as a binary variable (0 = non-smoker, 1 = current smoker) indicating active tobacco use. In all cohorts, weight and length were measured at the research center. Subsequently, body mass index (BMI) was calculated using the established formula weight(kg)/[length(m)]^2^. In the Rotterdam Study and UK Biobank, information on cell counts was also available and measured using the percentages of monocytes and lymphocytes.

#### Ascertainment of secondary outcomes

Having high blood pressure (no = 0, yes = 1) was defined in the UK Biobank by answering “High blood pressure” as one of the answers to the touch screen question: “Has a doctor ever told you that you have had any of the following conditions? (You can select more than one answer)”. In the Rotterdam Study and Leiden Longevity Study, high blood pressure was defined as a resting blood pressure exceeding 140/90 mmHg or the use of blood pressure lowering medication. Prevalent cardiovascular disease was defined as a diagnosis prior to blood sampling with a disease with ICD-10 code I20-I25, I48, I50, I60-64, I70, I73, or I48 in the UK Biobank; in the Leiden Longevity Study as I20, I21, or I60–I69, and the Rotterdam Study classification has been described more in depth elsewhere^47^. Incident cardiovascular disease was defined as a first diagnosis with any of these ICD-10 codes after blood sampling and before the end of follow-up: 31^*st*^ November 2020 in the UK Biobank, 1^*st*^ January 2020 in the Rotterdam Study, and 1^*st*^ November 2021 in the Leiden Longevity Study. Participants with prevalent cardiovascular disease were excluded from the incident cardiovascular disease analyses. Maximum hand grip strength was defined as the strength measured using a Jamar hand dynamometer across the two arms in the UK Biobank, the dominant hand in the Leiden Longevity Study, and the non-dominant hand in the Rotterdam Study. Self-reported poor health (no = 0, yes = 1) in the UK Biobank was defined as answering “Poor” or “Fair” to the question “In general how would you rate your overall health?” in the UK Biobank. When a person in the Rotterdam study answered “worse” on the question “How do you rate your health at this moment, compared to your peers” they were seen as having self-rated poor health. In the Leiden Longevity study, answering “Poor” or “Very poor” to “How would you rate your health in general?” was determined as self rated poor health.

### 4.4 Biomarker models

#### Linear

ElasticNet regression models were used to construct the linear models. This approach combines Lasso and Ridge regression, allowing for both variable selection and regularization. The loss function that is minimized is:

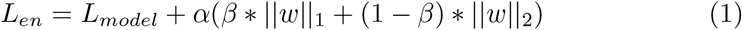

where *α* and *β* are regularization hyperparameters used to weigh the *L*_1_ and *L*_2_ loss, and *L*_*model*_ depends on whether the model is trained on the frailty index 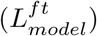 or all-cause mortality 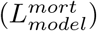. In the case of frailty index prediction, constitutes of the mean squared error loss:

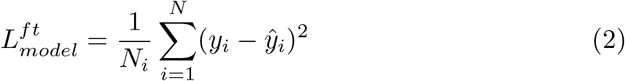

with *N* the training set size, *y*_*i*_ the frailty index value for participant *i*, and *ŷ*_*i*_ the estimated frailty index, which is a linear combination of the input: 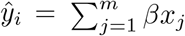 with *m* the amount of proteins, *β*_*j*_ the scalar pertaining to protein *j*, and *x*_*j*_ being the protein value of protein *j*. In the case of mortality prediction, *L*_*model*_ is defined by the cox regression loss:

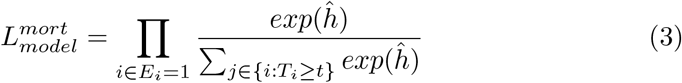

with {*E*_*i*_ = 1} the set of participants with an event, and {*i*: *T*_*i*_ ≥ *t*} the set of participants still at risk at time *t. ĥ* is the estimated hazard, defined by the linear combination of proteins as defined by the ElasticNet model.

In the training set, the linear models for the frailty index were built using sklearn”s ElasticNetCV v1.3.2. function and the mortality models were developed using sksurv”s CoxnetSurvivalAnalysis v0.22.2 with age at censoring as time variable and a binary variable indicating whether the participant was dead at the end of follow-up as event variable.

For both frailty index and mortality prediction, hyperparameter optimization was conducted using 5-fold cross-validation on the training set. A grid search over the *β* (ranging from 0.1 to 1 in steps of 0.01) was performed. For each *β*, a list of *α* values was automatically generated and evaluated to identify the optimal combination of *β* and *α*. For both prediction outcomes, the *α* values were logarithmically spaced between a data-driven maximum *α*_*max*_ and a minimum *α*_*min*_, where *α*_*max*_ is defined as the smallest value for which all coefficients are zero, based on the input data^48^: 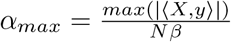. We consistently used the default settings for *α*_*min*_, corresponding to *α*_*max*_ × 10^−3^ for frailty predictions and *α*_*max*_ × 10^−4^ for mortality predictions. Subsequently, the performance of the resulting biomarker models was assessed using mean squared error for frailty index models and concordance index for mortality models. The best model was selected based on lowest error or highest concordance, respectively.

To avoid data leakage, this optimization was nested within the training set (so not touching the test or validation set). The test set was used for the selection of the final models based on performance metrics, while the validation set remained untouched during all modeling decisions. After model selection on the test set, final models were retrained on the combined train and test set (90%) using the previously selected hyperparameters in the nested training on the train set, and evaluated on the held-out validation set (10%) to report final performance and biomarker values.

#### Deep Learning

The deep learning models consist of a sequence of alternating linear layers with dropout and reLU nonlinearities. Due to the difference in input dimensionality, slightly different architectures were used for the different protein sets used; See Supplementary Table 9 for architecture details. All architectures end in a single output neuron, which represents the estimated frailty or log-risk function when trained on the frailty index or mortality, respectively.

The same architectures were used regardless of whether the model was trained on the frailty index or mortality, except for having one extra input neuron when training on frailty index to give age as an input, to match the ElasticNet input. Training on the frailty index uses a regularized mean squared error loss:

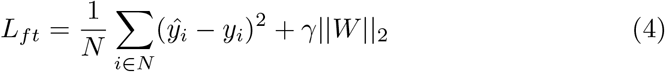

with *ŷ*_*i*_ and *y*_*i*_ being the estimated and true frailty index scores of participant *i*, and *N* being the number of training samples. *γ* is the regularization parameter, with ||*W* ||_2_ the *l*_2_-norm of the model parameters (i.e., weights and biases).

For training a FNN on mortality, following the approach from DeepSurv^49^, the standard cox partial likelihood loss was transformed into the average negative log partial likelihood:

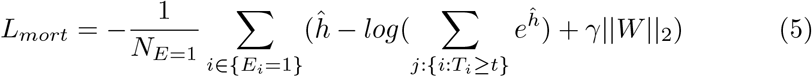

with *N*_*E*=1_ the number of participants with an observed event, *ĥ* the estimated negative log partial likelihood (i.e., the output of the neural network), {*i*: *T*_*i*_ ≥ *t*} being all the participants still at risk at time *T*_*i*_, and *γ*||*W* ||_2_ being the same regularization as in equation 4. All models were implemented using pyTorch.

We used a gridsearch approach to optimize the batch size, learning rate and regularization (*γ* in equations 4 and 5) parameters. For the models trained on frailty the gridsearch spanned from 8 to 32, 0.001 to 1.0 ∗ 10^−5^, and 0.01 to 1.0 ∗ 10^−4^ for batch size, learning rate and *γ*, respectively. For the mortality models, the gridsearch was conducted from 1000 to 20000, 0.001 to 1 ∗ 10^−6^, and 0.001 to 1.0 ∗ 10^−5^ for batch size, learning rate and *γ*, respectively. Final model parameters can be found in Tables 2 and 3.

**Table 2:** Hyperparameters for the ElasticNet models.

|  | Frailty index |  | All-cause mortality |  |
| --- | --- | --- | --- | --- |
| | $\alpha$ | $\beta$ | $\alpha$ | $\beta$ |
| Allprot | $2.5 * 10^{-4}$ | 0.99 | $1.6 * 10^{-3}$ | 0.99 |
| CMB | $1.5 * 10^{-4}$ | 0.99 | $8.4 * 10^{-3}$ | 0.1 |
| FFS | $2.2 * 10^{-5}$ | 0.99 | $7.6 * 10^{-4}$ | 0.69 |

**Table 3:**
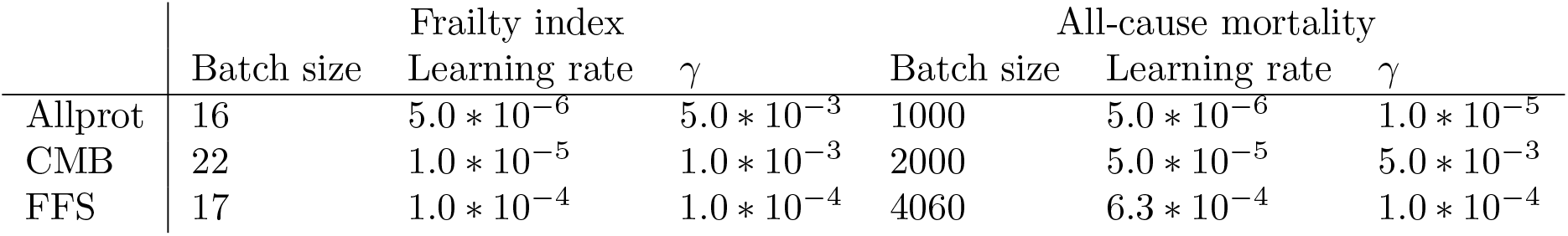
Model hyperparameters for the Feedforward Neural Networks.

While ElasticNet models were optimized using cross-validation, this approach was infeasible for the deep learning models due to the computational complexity. Therefore, the deep learning models were optimized by picking the hyperparameter combination with the highest result on the test (20% hold-out) set (leaving the validation set untouched).

Afterwards, our 70/20/10 data split was transformed into a 90/10 data split, retaining the same 10% hold-out validation set and merging the train and test sets. The final models were trained and evaluated on the validation set with the previously determined model parameters. 10 different seeds for parameter initialization were run, and the best model was selected based on either *R*^2^ score or C-index (for the frailty index and all-cause mortality, respectively) in the 90 % training set, which was then used for association analysis in the validation set.

### 4.5 Forward Feature Selection

For implementing the forward feature selection procedure, we used the SequentialFeatureSelector function from SKLearn v1.3.2. In short, all features are used to independently create a univariate model to estimate the frailty index (or allcause mortality). Next, these models are tested using the R2 score (or C-index). Using a 5-fold nested cross-validation setup on the training set, the protein with the best score metric is selected as the first (most important) protein. In the subsequent rounds, all linear combinations with the previously selected protein(s) are tested using linear regression, and the best scoring proteins are selected. This process repeats until the scoring metric does not improve more than the tolerance parameter, which was set at 0.001. Note that this procedure only uses the training set, leaving the test and validation set untouched. This resulted in 27 proteins and age selected for the frailty index and 20 for mortality; see Supplementary Table 4 for the specific proteins.

### 4.6 Statistical model comparison

To compare the performance of different modeling choices for the aging biomarkers, we applied bootstrap resampling (100 iterations) after completing hyperparameter optimization (on the train (EN) or train and test (FNN) data). This procedure was designed solely to quantify the variability of goodness-of-fit estimates (e.g. R^2^ or C-index), not to further optimize the biomarkers. In each iteration, the hyperparameters selected on the original training set were reused. We drew a bootstrap resample from the 90% combined training and test set (sampling participants with replacement) and evaluated model performance on the fixed 10% validation set. Next, to determine whether goodness-of-fit metrics differed significantly between sets, we first assessed the normality of differences using the Shapiro-Wilk test (shapiro function, SciPy v1.12.0). Since all p-values were greater than 0.05, indicating that the differences followed a normal distribution, we proceeded with a paired Z-test assuming unequal variance (ztest function, StatsModels). P-values were false discovery rate adjusted using the Benjamini-Hochberg procedure^50^. A false discovery rate p-value threshold of <0.05 was used to define statistical significance.

### 4.7 Calculation of previously developed aging biomarkers

We calculated PAC following the dedicated R-script^32^. The ProteinMortality Score and MetaboMortality Score from Gadd et al.^31^ were calculated by multiplying the included proteins and metabolites by their respective weights and calculating a sum score. MetaboHealth was calculated using the MiMIR package^51^. In the Rotterdam Study, we calculated DNAm Hannum, DNAm Horvath using the methylclock R-package^52^ and DNAm GrimAge using the Python scripts provided by the researchers who developed these biomarkers.

### 4.8 Association Analysis

In order to determine the component of each biomarker independent of age, we created a linear regression predicting chronological age using the biomarker score for each biomarker^53^ on the validation set. Then, we determined the residuals of each of these linear regressions, which indicate whether an individual is biologically “older” or “younger” than the population average for their chronological age. After calculating the residuals, to enhance comparison among aging biomarkers, we applied a Z-transformation to these residuals and used this Z-transformed residual in the subsequent analyses with end-points.

Linear regression analysis using the OLS function from statsmodels v0.14.1 with an added constant was used to assess the associations of the aging biomarkers with continuous outcomes. In case of BMI and smoking, the aging biomarkers were used as the dependent variables in this analysis, as lifestyle factors are hypothesized to influence biological aging, and aging biomarkers are intended to reflect that. Associations of aging biomarkers with binary outcome measures were assessed with logistic regression analyses using the Logit function from statsmodels v0.14.1 with an added constant. Additionally, associations of aging biomarkers with incident outcomes were assessed using Cox-Proportional Haz-ard models using the CoxPHFitter function from lifelines v0.29.0.

In all analyses, the first model included only sex as a covariate. In the second model, socioeconomic status was additionally added as a covariate. In the third model, next to the covariates from the previous models, BMI and smoking were added as a covariate. In the Rotterdam Study and UK Biobank, we performed analyses in a fourth model and additionally adjusted the analyses for cell counts. Next, we performed the analyses adjusting for the fourth, or in the Leiden Longevity Study third, model with additionally either GDF15 or MetaboHealth to determine whether the associations were independent of these previously established biomarkers. Z-tests were used to assess whether effect sizes between aging biomarkers differed statistically significantly.

## 5 Supplementary information

### 5.1 Frailty index deficits

#### UK Biobank

**Table 1:**
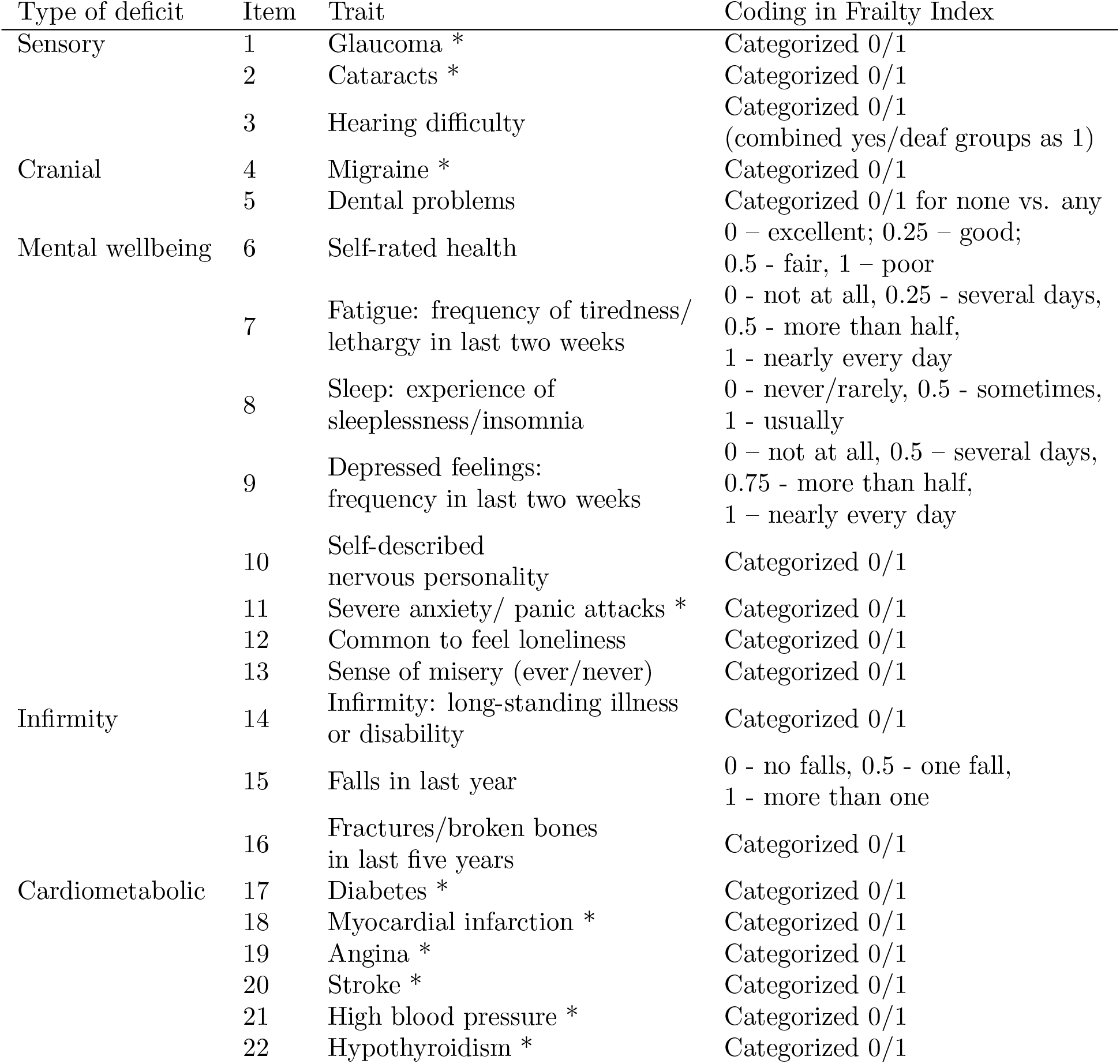
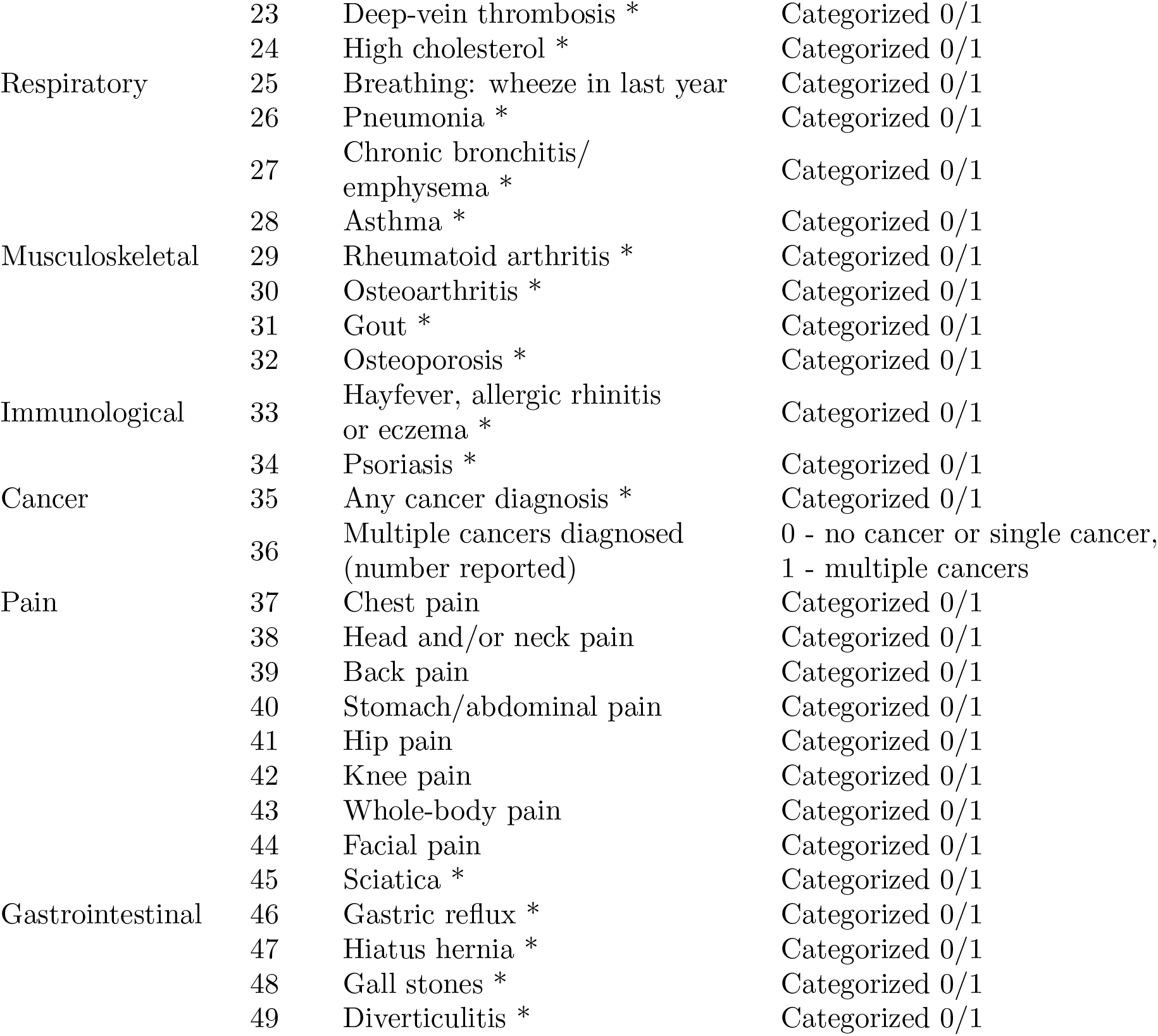
The Rockwood frailty index in the UK Biobank, as developed by Williams et al. [25], following the procedure of Searle et al. [54]. The Rockwood frailty index in the UK Biobank consist of 49 items, coded according to the table above. The frailty score is calculated by summing all 49 codes and dividing by the total number of items.

**Table 2:** Placeholder for frailty deficits in the Rotterdam Study (too big to fit on page)

### 5.2 Multi-omics

We explored several methods for integrating metabolomics and proteomics for biomarker training: concatenate, principal component analysis (PCA) and then concatenate, combining with MetaboHealth and Angle-Based Joint and Individual Variation Explained (AJIVE). For the concatenate method, we simply concatenated all proteins and metabolites together. For the PCA and then concatenate method, we first took separate PCAs for the protein and metabolomics data, then concatenated the first 10 or 20 principal components. When combining with MetaboHealth, we added the MetaboHealth score[11] to all of the protein features. Finally, AJIVE is a method for identifying shared and separate components from two data modalities [55] [56]. Using AJIVE, we identified the shared and individual components of the metabolomics and proteomics data, and used those to train our biomarkers.

Supplementary Table 3 shows that none of the aforementioned methods increase either the *R*^2^ score or the C-index when trained on the frailty index or mortality, respectively, compared to the baseline using only the cardiometabolic proteins (protein-only). However, this protein-only model was trained on a larger set of participants, due to reduced metabolomics availability. When reducing the training set size for the protein-only model, some deterioration in performance can be seen, which can partially be compensated for by adding metabolites at the input (through concatenation). However, adding more complex multi-omics approaches did not seem to improve performance further.

**Table 3:** Goodness-of-fit metrics on various multi-omics integration methods.

| Dataset / method | ElasticNet |  | Feedforward Neural Networks |  |
| --- | --- | --- | --- | --- |
|  | Frailty index (R2) | All-cause mortality (C) | Frailty index (R2) | All-cause mortality (C) |
| Using only cardiometabolic proteins | 0.24 | 0.71 | 0.25 | 0.71 |
| Cardiometabolic proteins in population subset with metabolomics | 0.23 | 0.69 | 0.20 | 0.68 |
| Cardiometabolic proteins and NMR-metabolites concatenated | 0.24 | 0.69 | 0.23 | 0.69 |
| Using AJIVE | 0.17 | 0.61 | 0.14 | 0.62 |
| PCA then concatenate, using 10 principal components | 0.17 | 0.61 | 0.16 | 0.64 |
| PCA then concatenate, using 20 principal components | 0.19 | 0.67 | 0.17 | 0.66 |
| Cardiometabolic proteins and MetaboHealth, concatenated | 0.23 | 0.69 | 0.23 | 0.68 |

### 5.3 Selected proteins in Forward Feature Selection

**Table 4:**
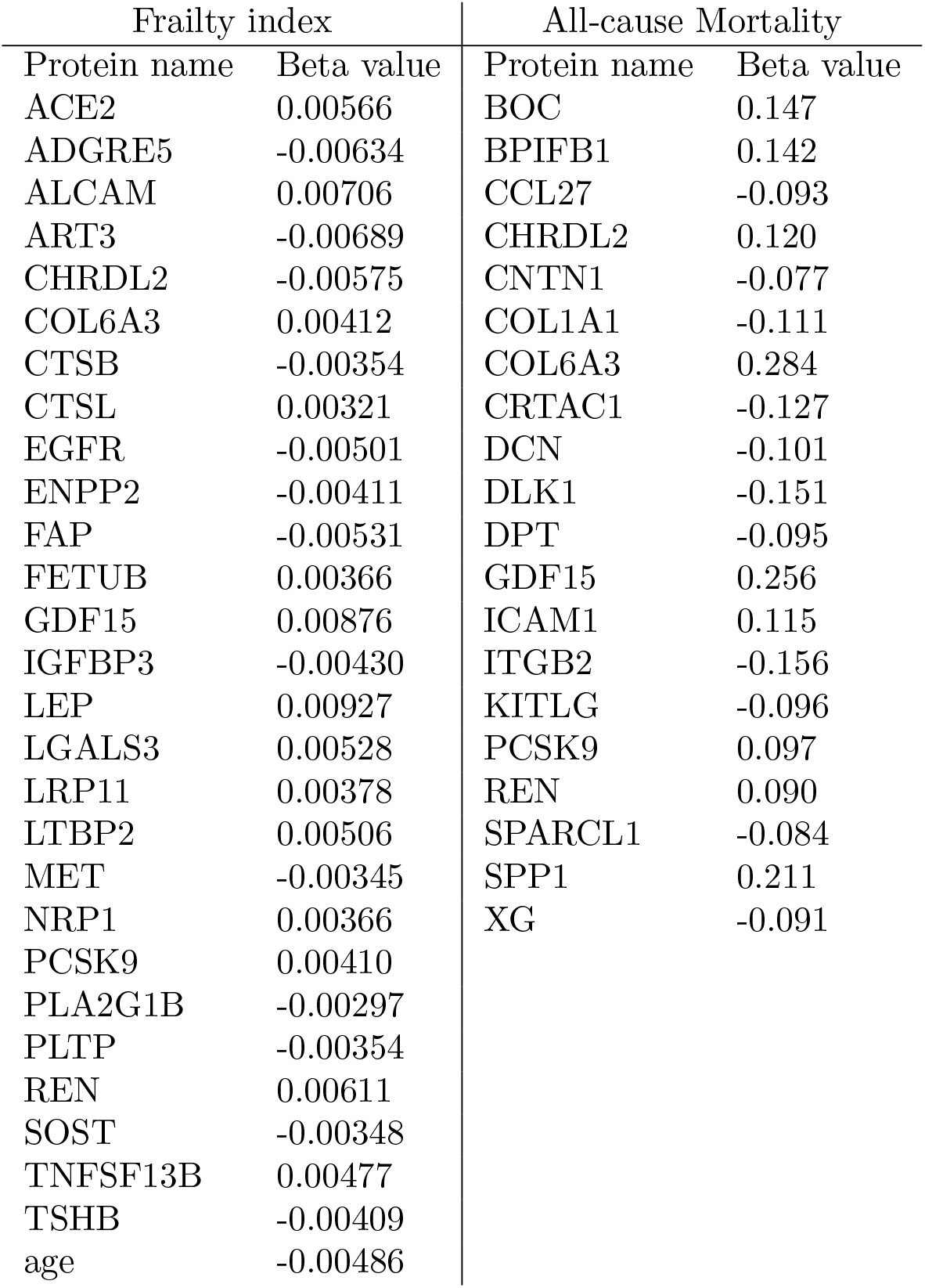
The proteins selected in the Forward Feature Selection procedure, together with the accompanying beta values of the linear regressor used for selection.

### 5.4 Full comparison between allprot, CMB and FFS

**Fig. 1:**
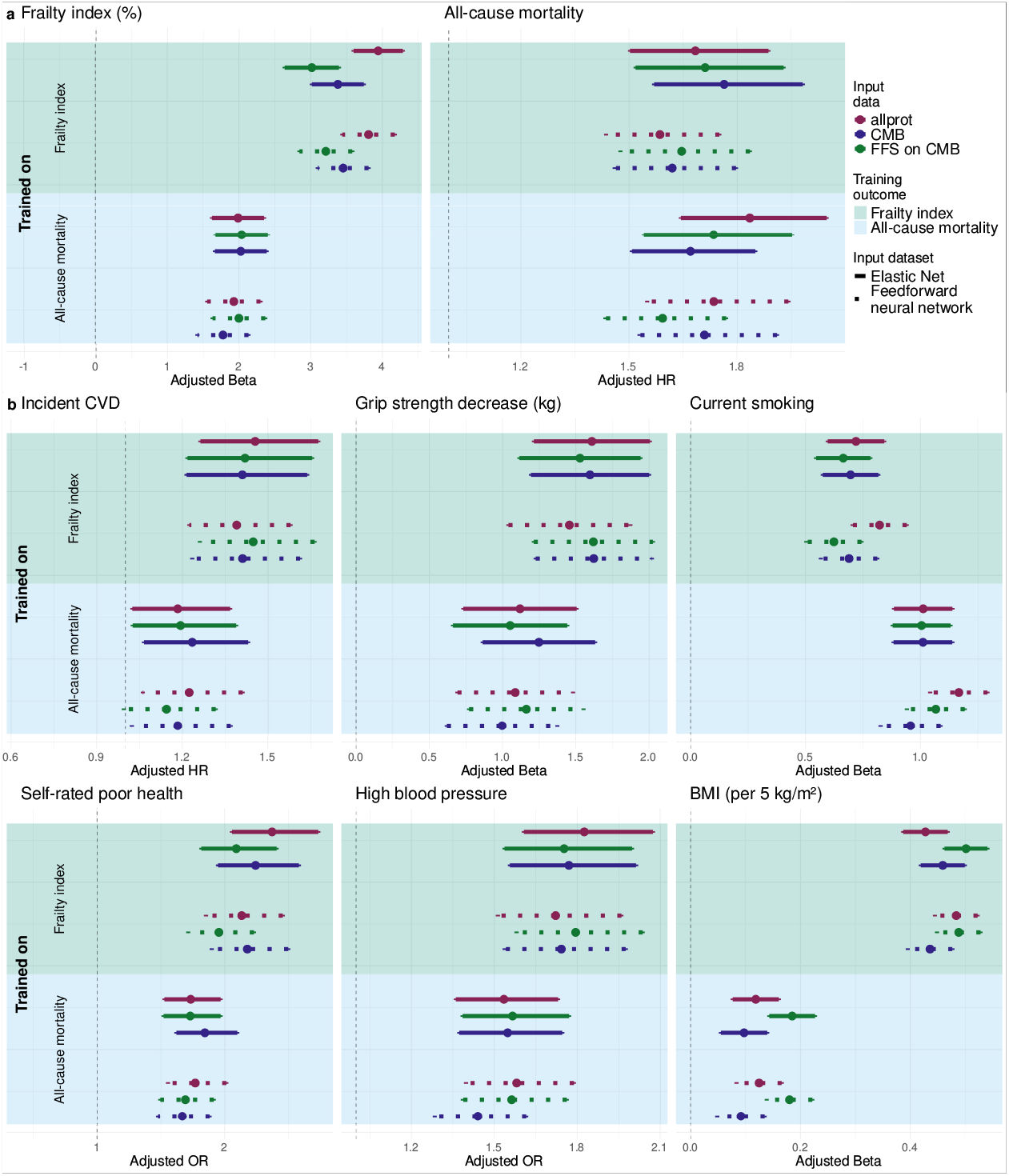
Full comparison on the training endpoints between biomarkers trained on the full set of 1428 proteins (allprot), the subset defined by the cardiometabolic panel (CMB), and the subset of CMB defined by the forward feature selection (FFS) procedure.

### 5.5 External validation of biomarkers

**Table 5:** placeholder for table with all point estimates (too big to fit on page).

**Fig. 2:**
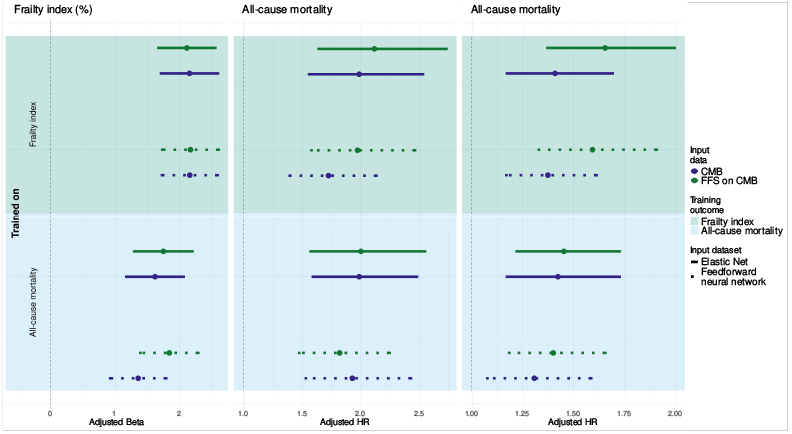
External validation of constructed biomarkers on the Rotterdam Study (left and middle column), and the Leiden Longevity Study (right column)

**Table 6:** placeholder for table with data from full RS cohort (too big to fit on page).

### 5.6 Data splits

**Fig. 3:**
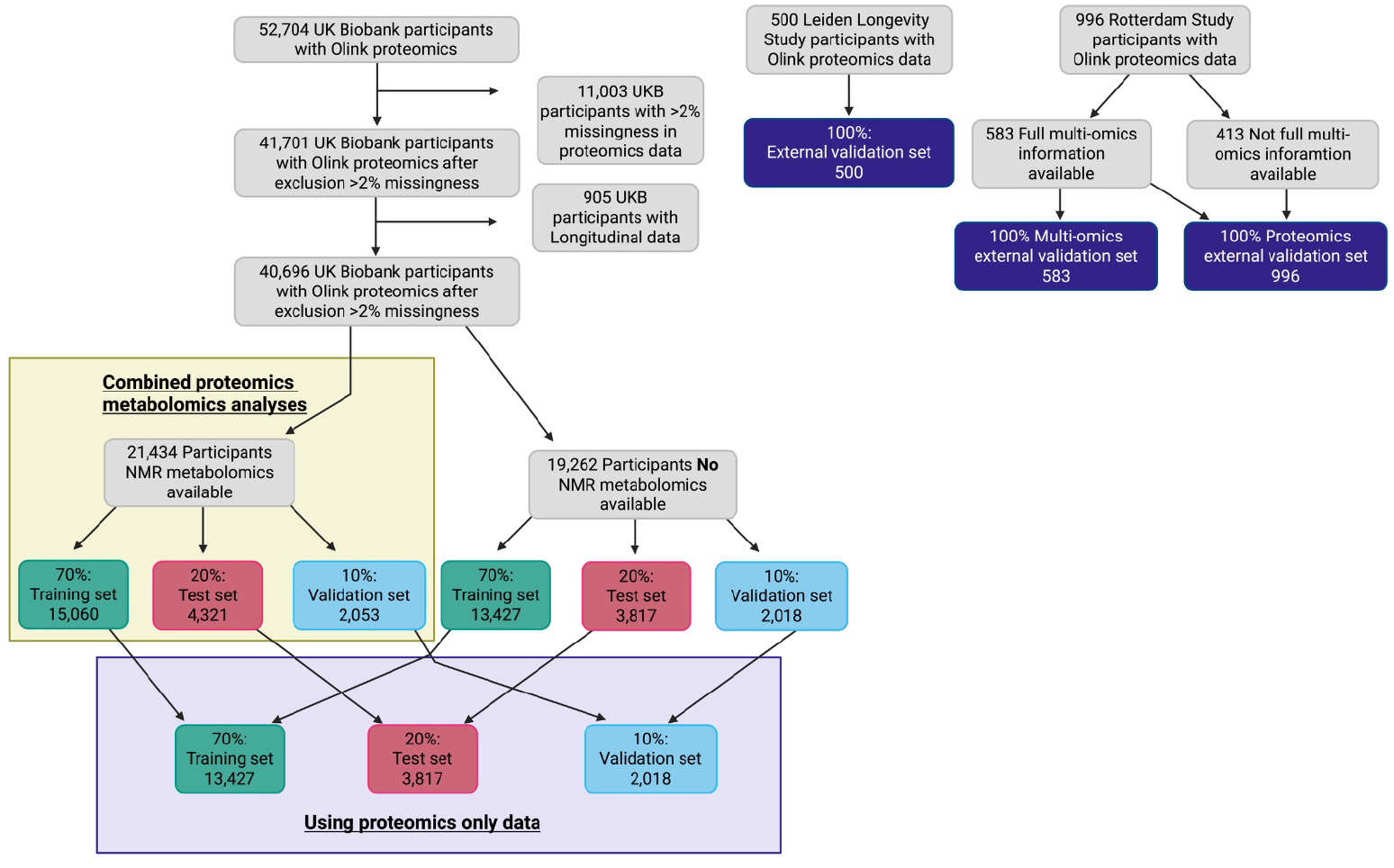
Flowchart of data split creation on the UK Biobank, Leiden Longevity Study and the Rotterdam Study

### 5.7 Sex-specific analysis

**Table 7:** Concordance indexes on the four protein panel (allprot) dataset using various adjustments for sex.

|  | Linear | Deep |
| --- | --- | --- |
| No adjustment | 0.73 | 0.73 |
| Sex as (input) covariate | 0.73 | 0.73 |
| Men only | 0.72 | 0.70 |
| Women only | 0.71 | 0.72 |

### 5.8 Highest correlations between proteins in the UK Biobank

**Table 8:** Placeholder for table with the highest correlating proteins for each protein in the UK Biobank.

### 5.9 Neural network architectures

**Table 9:** Neural network architectures.

|  | Layer 1 | Layer 2 | Layer 3 | Layer 4 |
| --- | --- | --- | --- | --- |
| Allprot | Linear: 1428* x 750 | Linear: 750 x 400 | Linear: 400 x 100 |  |
|  | Dropout: p = 0.2 | Dropout: p = 0.1 | Dropout: p = 0.1 | Linear: 100 x 1 |
|  | ReLU | ReLU | ReLU |  |
| CMB | Linear: 344* x 200 | Linear: 200 x 100 | Linear: 100 x 10 |  |
|  | Dropout: p = 0.2 | Dropout: 0.1 | ReLU | Linear: 10 x 1 |
|  | ReLU | ReLU |  |  |
| FFS | Linear: 20-28** x 15 | Linear: 15 x 10 | Linear: 10 x 5 |  |
|  | ReLU | ReLU | ReLU | Linear: 5 x 1 |

\* input dimension is 1 higher when trained on the frailty index, to give age as an input \*\* input dimension is 20 for mortality-trained, and 28 for frailty index-trained models.

